# Different time scales used for sexual partner surveys pose a challenge in modelling dynamics of sexually transmitted infections

**DOI:** 10.1101/2023.12.25.23300526

**Authors:** Hiroaki Murayama, Akihiro Nishi, Akira Endo

## Abstract

Mathematical models for sexually transmitted infections (STIs) are parameterised by empirical data on sexual behaviour (e.g. the number of partners over a given period) obtained from surveys. However, the time window for reporting sexual partnerships may vary between surveys and how data for different windows can be translated from one to another remains an open question. To highlight this issue, we compared the distributions of the number of sexual partners over one year and four weeks from the British National Surveys of Sexual Attitudes and Lifestyles. The results show that simple linear rescaling did not render the one-year and four-week partner distributions aligned. Parameterising STI models using survey-based sexual encounter rates without considering the implication of the reporting window used can lead to misleading results.

## Main text

Sexually transmitted infections (STIs) have been a major global health concern with a substantial disease burden *(1, 2)*. In addition to conventional STIs such as syphilis, gonorrhoea, trichomoniasis and chlamydia, STIs can also emerge in the form of an outbreak such as human immunodeficiency virus (HIV) *(3)* and mpox (which spread through sexual contacts in the recent outbreak, if not formally established as an STI) *(4)*. One of the key factors to the establishment and maintenance of STIs in populations is the heterogeneity in sexual behaviours and network structures, where relatively few individuals with many more sexual partners than average play a crucial role in shaping transmission dynamics *(5–9)*. Individuals exhibit diverse sexual partnerships not only in quantity (i.e. number) but also in quality, e.g. either casual or steady (long-term) relationships *(10–21)*. Causal partnerships tend to form and break frequently and thus contribute more to the number of partners over a given period of time *(16–18)*. Similarly, concurrent partnerships or contacts between commercial sex workers and their clients can greatly increase one’s rate of sexual encounters; they may also contribute to the formation of dense networks *(19, 20)*. Individuals with high rates of sexual encounters are at higher risk of both infection and transmission and are often referred to as the “core group” to reach out with STI prevention efforts. Understanding how STIs are transmitted over the sexual network among the core groups and beyond is critical to developing the most effective approaches to disease control, for which mathematical models have been proven a powerful tool *(22)*.

Mathematical models have represented the dynamics of STIs through networks using empirical datasets on sexual behaviour. In particular, network models are used to represent static or dynamic sexual relationships between individuals in order to simulate and/or infer STI spread patterns in the population. Such network models are often parameterised by the reported number of sexual partners or sexual activities over defined time windows in questionnaire-based surveys *(6, 23, 24)*.

However, the time windows for reporting sexual behaviours often vary between sexual partner surveys *(25–29)*. Typical windows include 1 month (or 4 weeks), 3 months, 1 year or lifetime. Methods for ensuring comparability and generalisability across data over different reporting windows have not been proposed or established. This poses problems for parameterisation of network models for STIs. The basic reproduction number R_0_, defined as the mean number of secondary transmissions caused by a typical infected case, is one of the key metrics that shapes the transmission dynamics of STIs. In sexual network models, it could be determined by the unique number of sexual partners of cases and/or the frequency of sexual acts between these partners over the infectious period (i.e. the duration of infectiousness of an infected individual) of the pathogen of interest *(6)*. Therefore, sexual behaviour over the duration of the infectious period is the most relevant data to inform transmission models, but in most cases, it is not readily available. The infectious period of STIs vary widely—from weeks (e.g. gonorrhoea and mpox) to decades (e.g. HIV and HPV)—but only a limited number of reporting windows could be used in sexual behaviour surveys for logistical reasons. Besides, it is sometimes more practical to use reporting windows that differ from the infectious period. For example, setting reporting windows that are too short can lead to loss of information because individuals may report no sexual activity for a short period of time regardless of their long-term sexual behaviour. For these reasons, parameterisation of mathematical models usually involves rescaling of empirical sexual behaviour data (e.g. by assuming a constant rate of sexual encounters), the potential implications of which have not been fully discussed.

In the present study, we compared (in a rather naive manner) the distribution of sexual partners over different reporting windows (i.e. 4 weeks and 1 year) from the same cohorts of British National Surveys of Sexual Attitudes and Lifestyles (Natsal; 2000 and 2010 cohorts combined) *(25)*. We fitted a Weibull distribution, left-truncated at 1, to each of the 4-week and 1-year male-to-male partner number data. The Weibull distribution has been shown in our previous study *(6)* to well represent the heavy-tailed nature of empirical sexual partnership data among men who have sex with men (MSM; one of the key population groups for STI prevention *(30)*). While the Weibull distribution is typically characterised by the shape parameter α and the scale parameter θ, here we used an alternative parameterisation: shape α and a Pareto-approximated exponent κ (κ=α/θα) for estimation stability *(6)*. The posterior median estimates were α=0.16 (95% credible interval [CrI]: 0.03, 0.35) and κ=1.32 (95% CrI: 1.08, 1.58) for 4-week partners and α=0.10 (95% CrI: 0.02, 0.19) and κ=0.77 (95% CrI: 0.66, 0.88) for 1-year partners. We then rescaled the distribution for 1-year partners to the 4-week window by linearly reducing the Weibull distribution by 13-fold (given by 365/28) and truncating it again at 1. This rescaling resulted in parameter values of α=0.10 (95% CrI: 0.02, 0.19) and κ=1,00 (95% CrI: 0.87, 1.16), which were significantly different from the parameters estimated from the 4-week partner data (i.e. the above-mentioned α=0.16 and κ=1.32) (Figure 1).

**Figure 1.**
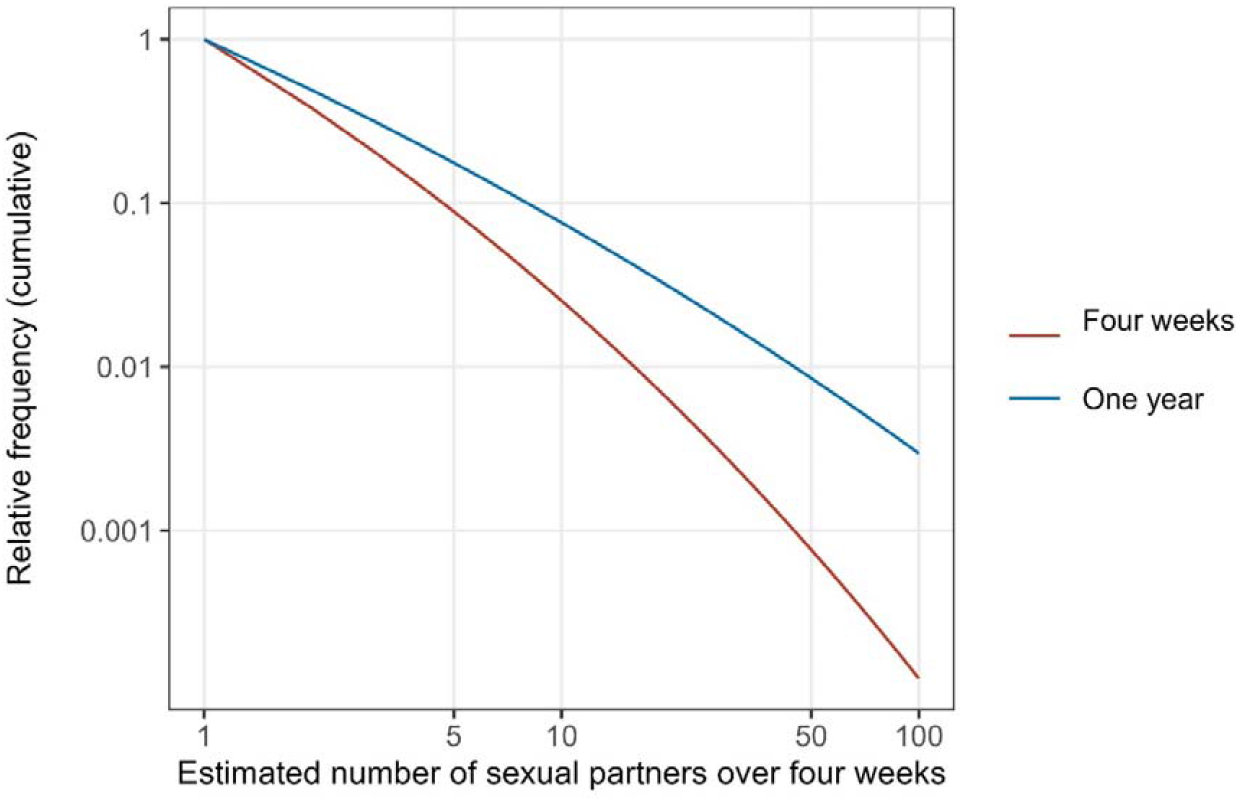
Comparison of the distributions of the number of male-to-male sexual partners over four weeks and one year from the British National Surveys of Sexual Attitudes and Lifestyles. The curves on a log-log plot show the left-truncated Weibull distributions fitted to the reported number of sexual partners. The distribution for the one-year partners was rescaled by a factor of 1/13 to be comparable to that for the four-week partners.

These results highlight the need for both theoretical and empirical studies on how to translate sexual behaviour data reported for one time window into another. Many STI modelling studies have used annual numbers of partners from surveys to specify the sexual contact rates assuming proportionality *(6, 7, 31)*. While this practice may have been unavoidable due to data availability, it may have been a naive, if not proven wrong, approach to parameterising models. Previous studies have often categorised sexual partners into different types of relationships, such as steady and casual partners, which are known to show different patterns of relationship duration *(10–13, 21)*. The algorithm that could translate between the numbers of partners over short- and long-term reporting windows thus would not be straightforward; in particular, individuals with many partners, who play a key role in STI dynamics *(6–9)*, may have a mixture of steady and casual partners. Future studies combining both theoretical and empirical approaches from infectious disease epidemiology and social sciences are warranted to improve our understanding of dynamic sexual behaviour and to achieve more plausible parameterisation of STI models.

## Acknowledgement

HM and AE are supported by Japan Science and Technology Agency (grant number JPMJPR22R3, to AE). AN and AE are supported by the National Institutes of Health (K01AI166347, to AN), the National Science Foundation (NSF #2230125, to AN) and the Japan Science and Technology Agency (JPMJPR21R8, to AN). AE is supported by Japan Society for the Promotion of Science (Overseas Research Fellowships; Grants-in-Aid *KAKENHI* 22K17329). The content is solely the responsibility of the authors and does not necessarily represent the official views of the funders. The funders had no role in study design, data collection and analysis, decision to publish, or preparation of the manuscript.

## Data availability

The underlying data (National Survey of Sexual Attitudes and Lifestyles, UK) is available from UK Data Service (serial numbers: SN 7799, and SN 8865) provided the End User License Agreement.

## Ethics

This study was approved by the London School of Hygiene & Tropical Medicine ethics committee (reference number: 27985).

